## Supplementary figures for "Genomic epidemiology and associated clinical outcomes of a SARS-CoV-2 outbreak in a general adult hospital in Quebec"

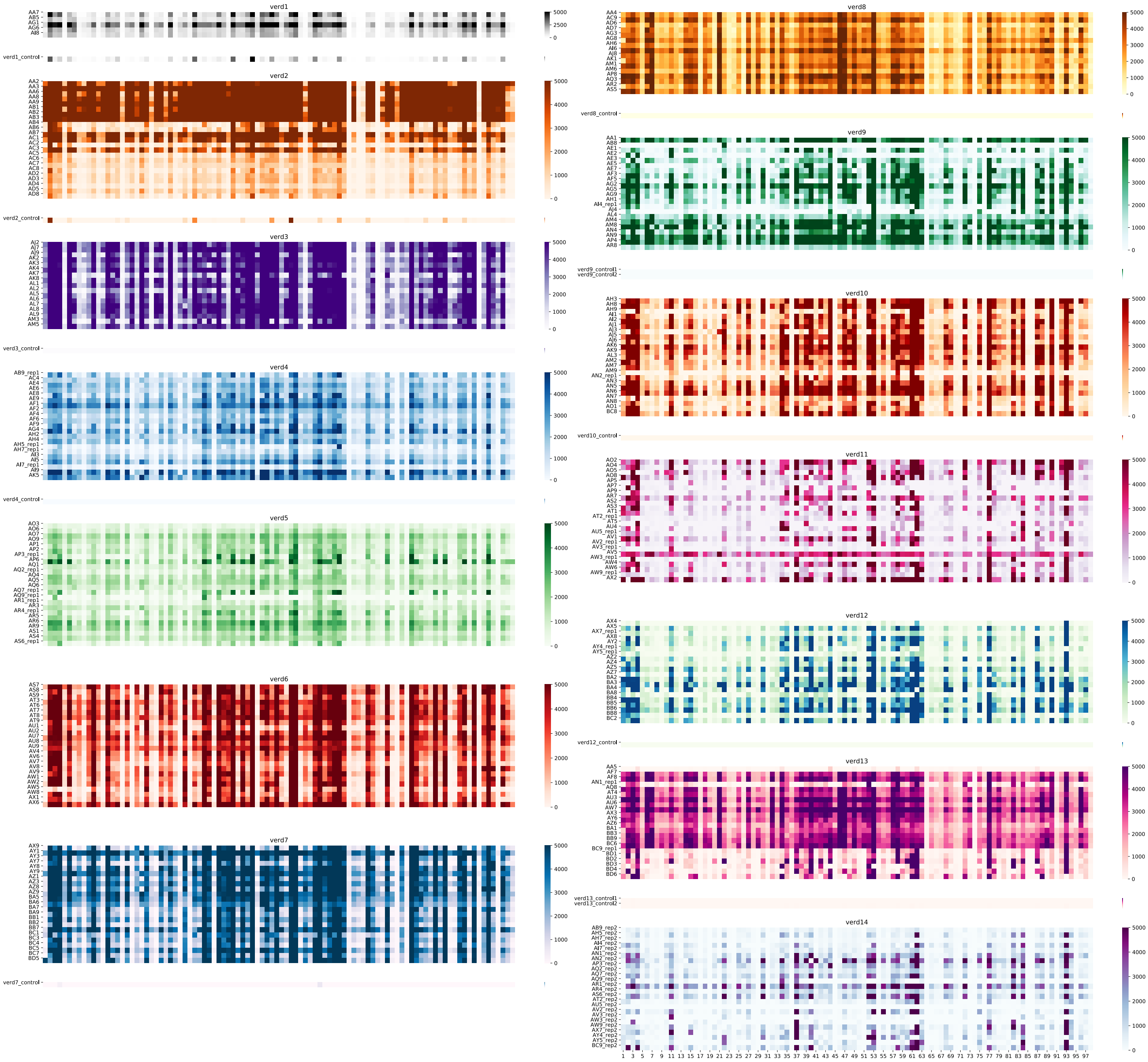


**Supplementary Figure 1: Amplicon coverage for all samples.**
Amplicon coverage for each sample, including the controls, was calculated using bedtools using a 90% overlap between the query and the target (ARTIC Network amplicon coordinates) as well as 80% overlap between the target and the query.

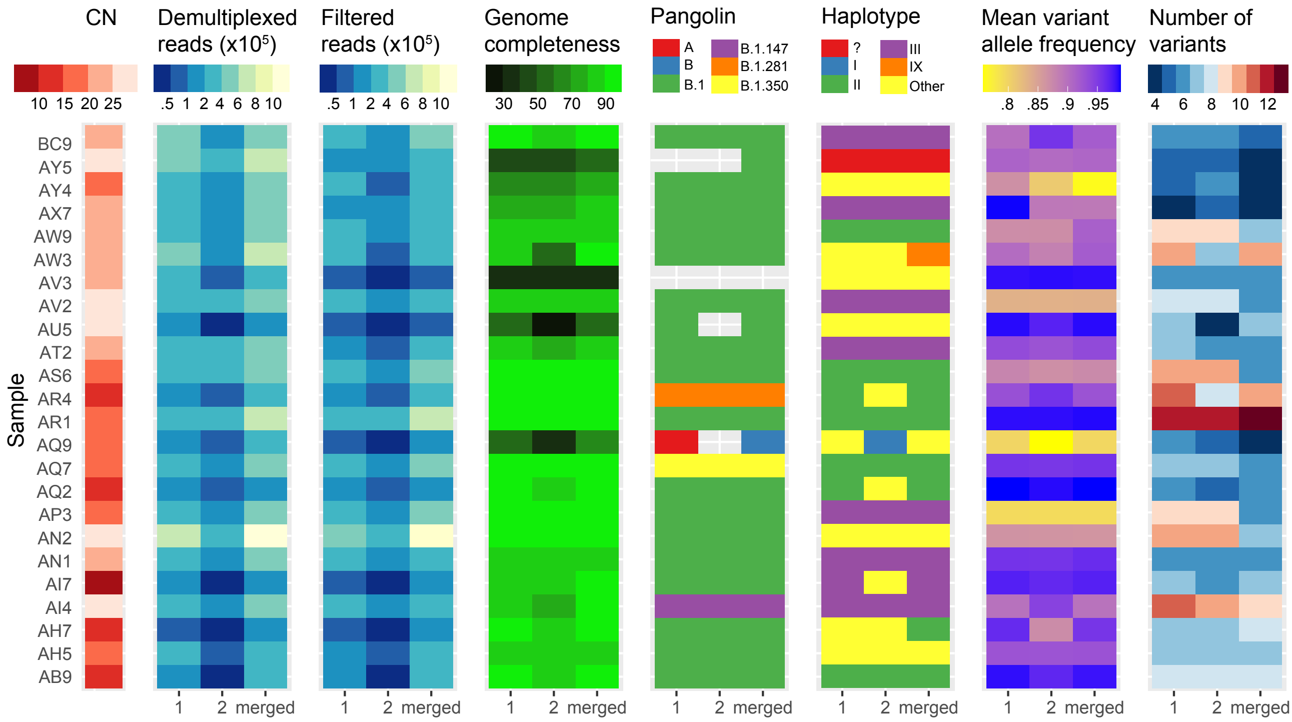


**Supplementary Figure 2: Technical replicates for a selection of samples.** Two separate nanopore sequencing library preparations (1 & 2) from the same PCR products and the corresponding merged data (merged) on the horizontal axis. Results generated from the Medaka version of the ARTIC Network bioinformatics SOP. Genotype data generated with modified consensus genomes that contain the most frequent variant ($> 50\%$) at any given position.

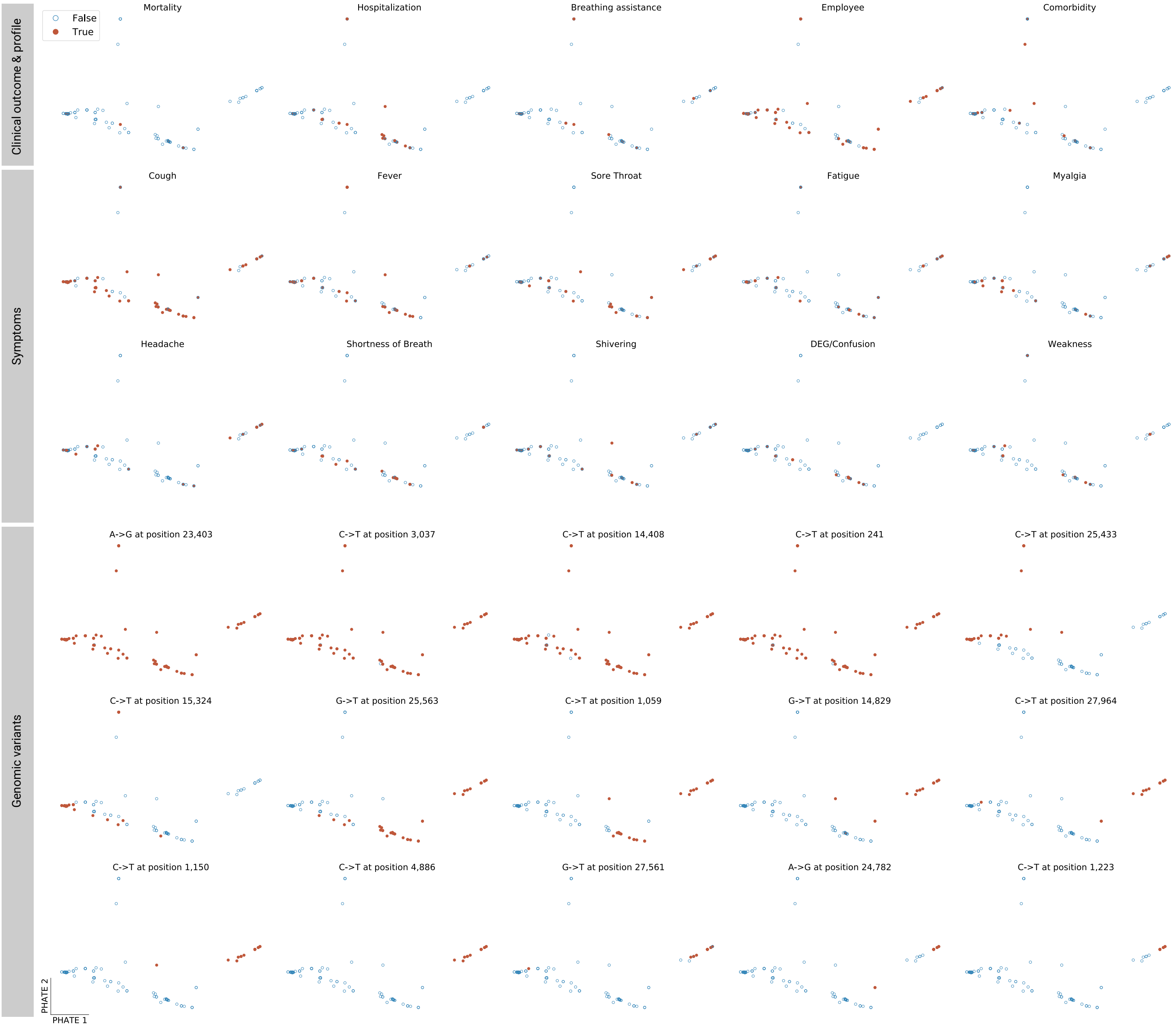


**Supplementary Figure 3: PHATE embedding of SARS-CoV-2 genomic variation.** Each point corresponds to one SARS-CoV-2 genome annotated with various clinical labels and symptoms. The 15 most frequent variants are annotated in the bottom panel. Sub-clade IIa (right cluster) is characterized by a number of variants differentiating it from sub-clade IIb (tip of the lower branch), see Figure 4, top left panel.

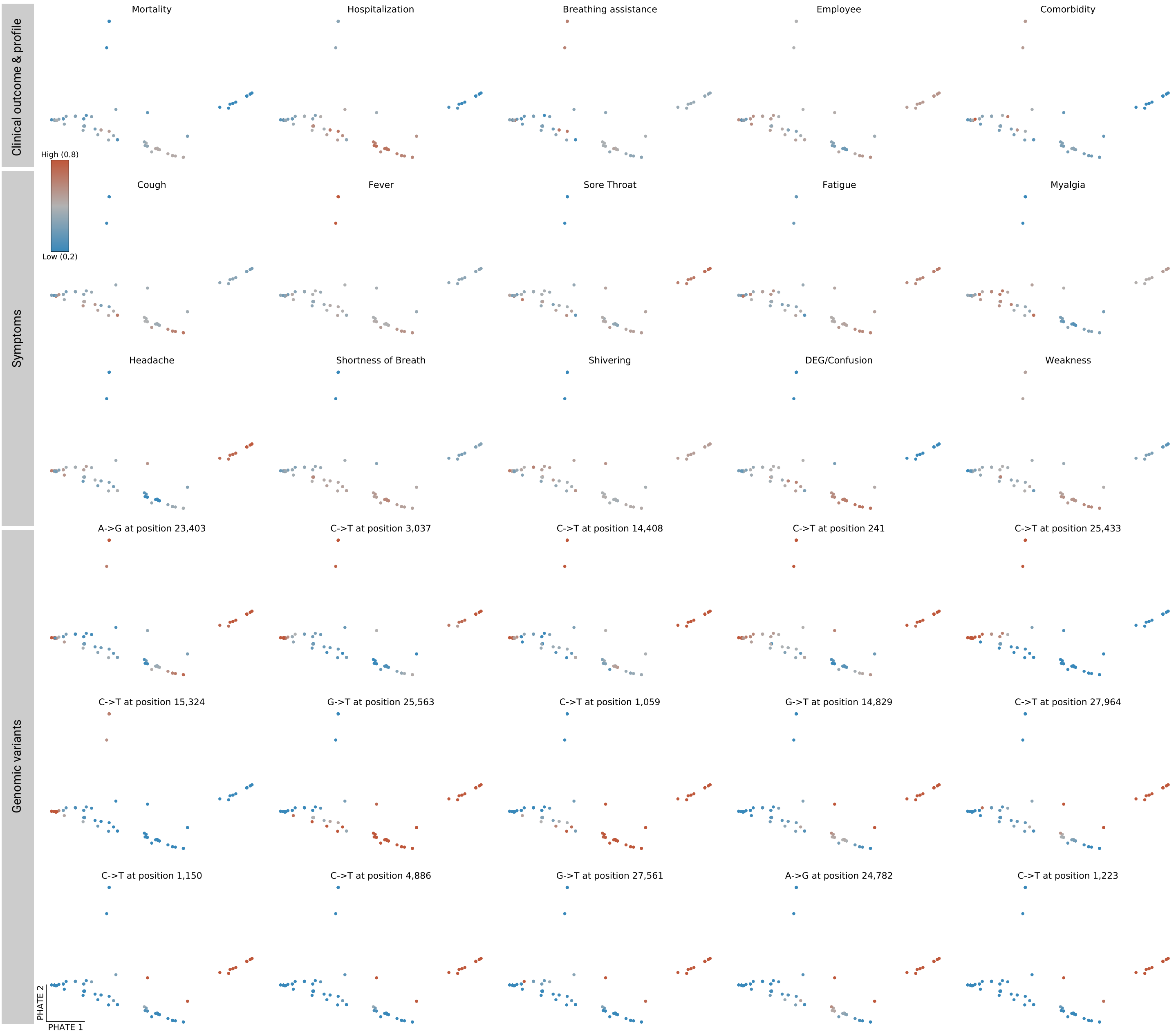


**Supplementary Figure 4: MELD relative likelihood estimates based on viral genomic variation.**

Likelihoods of various clinical labels and symptoms are displayed over the PHATE embedding of SARS-CoV-2 genomes (see **Supplementary Figure 3**). The subclade IIa cluster (top right right) appears depleted in adverse outcomes (mortality and hospitalization) and enriched in flu-like symptoms (sore throat, fatigue and headache). Conversely, the sub-clade IIb region (tip of the lower-right cluster) is slightly enriched in hospitalized patients and shows a higher likelihood of DEG/Confusion. MELD likelihoods of the 15 most frequent variants are displayed over the PHATE embedding of the genomes in the bottom panels. It should be stressed that MELD computes a local relative likelihood. Some manifold regions may be relatively depleted in a specific variant compared to other regions even if said variant is frequent in absolute terms, particularly in the case of 23403A¿G, 3037C¿T and 14408C¿T.

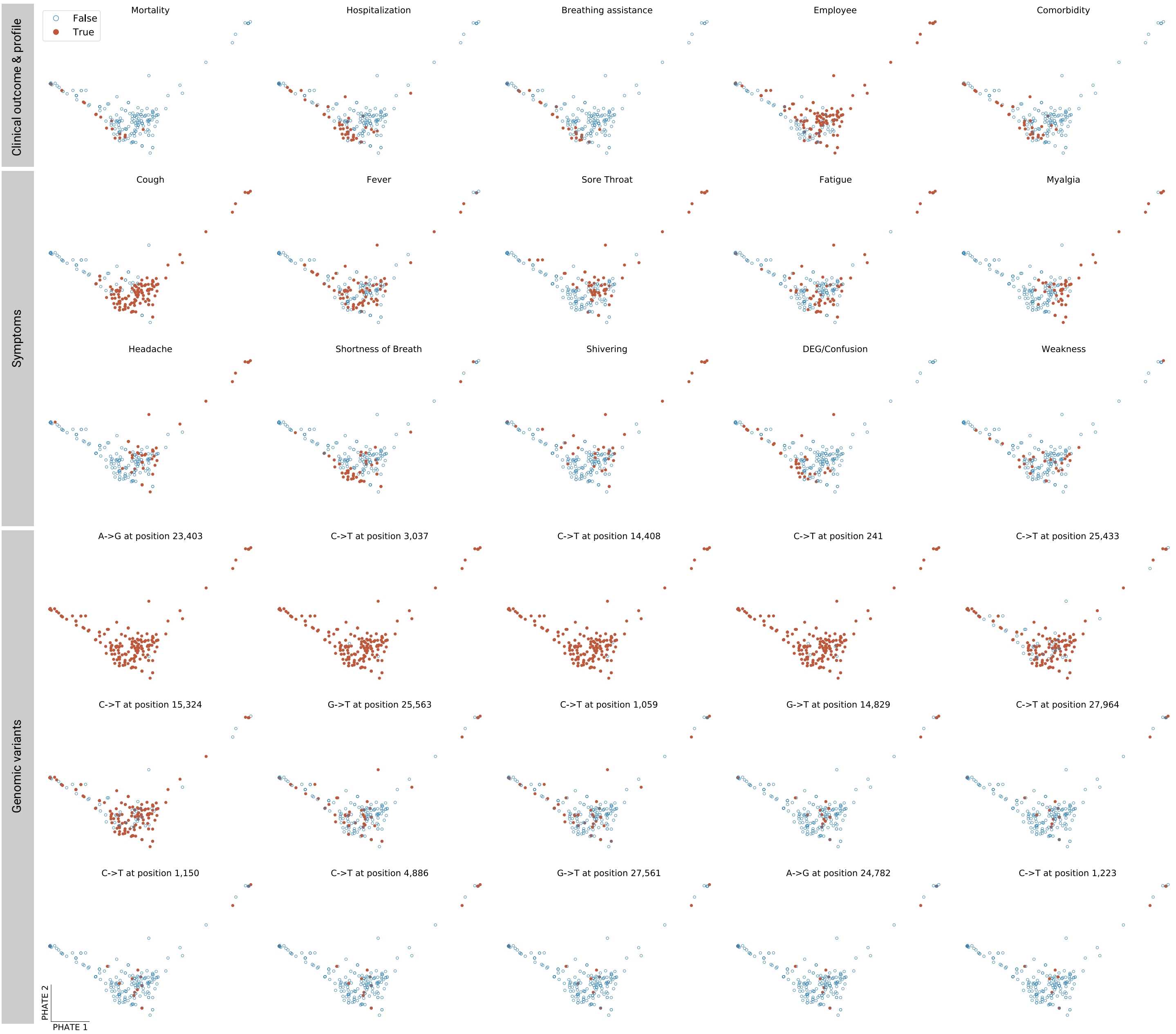


**Supplementary Figure 5: PHATE embedding of clinical features.**
Each point corresponds to one patient sample annotated with various clinical labels and symptoms (N.B. some samples correspond to the same patient, see Longitudinal Sequencing section ). The 15 most frequent variants are annotated in the bottom panel. Sub-clade IIa (right cluster) is characterized by a number of variants differentiating it from sub-clade IIb (tip of the lower branch), see Figure 4, top left panel.

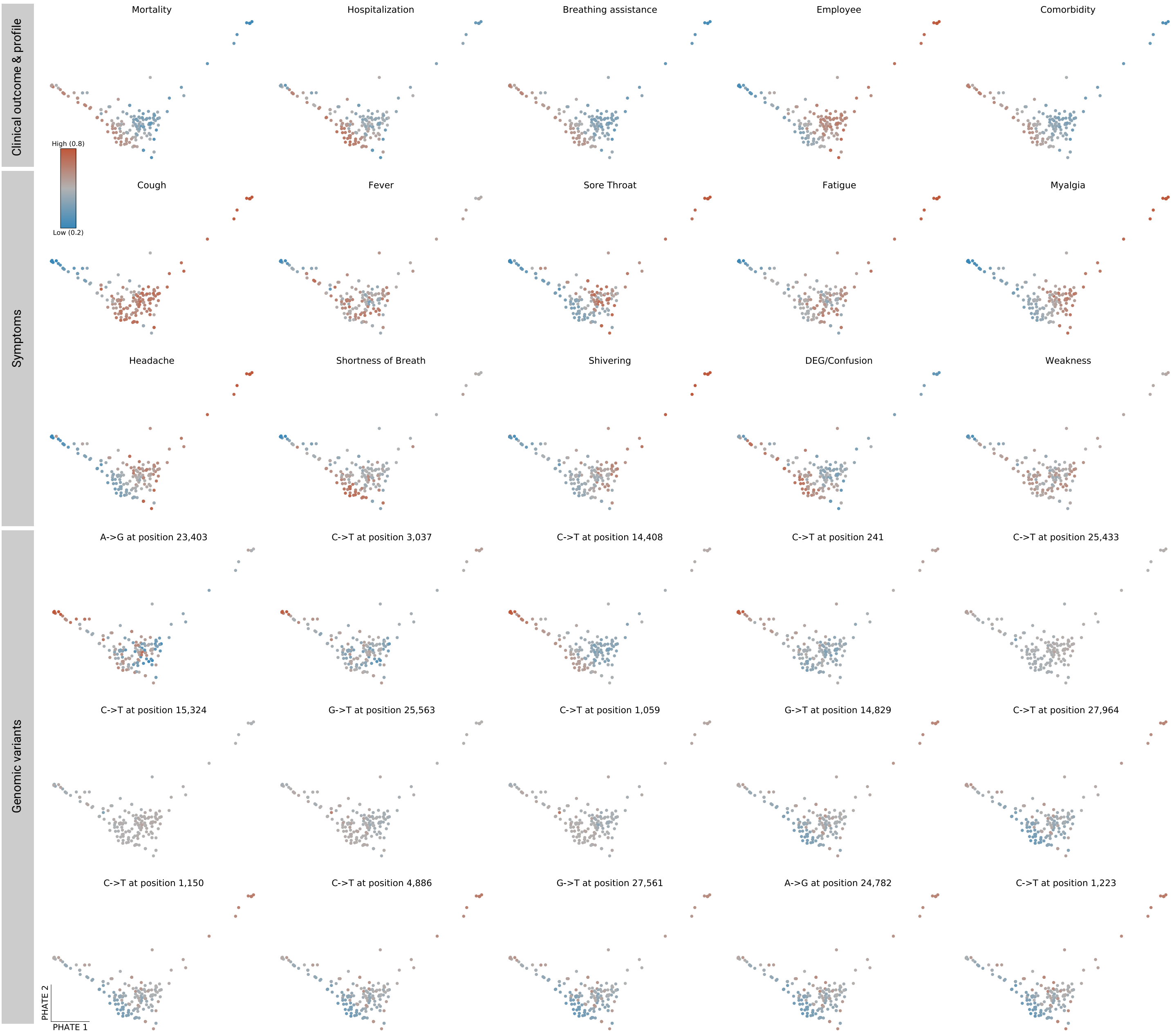


**Supplementary Figure 6: MELD relative likelihood estimates based on clinical features.** Likelihood estimates of various clinical labels and symptoms displayed over the PHATE embedding of the clinical features (see **Supplementary Figure 5**). The likelihood gradients of adverse outcomes (mortality, hospitalization and breathing assistance) are well aligned with comorbidity and DEG/Confusion gradients. Moreover, adverse outcome likelihoods appear to be inversely correlated with employee status as well as a set of flu-like symptoms (sore throat, myalgia, fatigue and headache). The MELD relative likelihood estimates of the 15 most frequent variants displayed over the PHATE embedding of the symptoms in the bottom panel.
